# Pharmacokinetic modelling to estimate intracellular favipiravir ribofuranosyl-5’-triphosphate exposure to support posology for SARS-CoV-2

**DOI:** 10.1101/2021.01.03.21249159

**Authors:** Henry Pertinez, Rajith KR Rajoli, Saye H Khoo, Andrew Owen

**Affiliations:** Department of Pharmacology and Therapeutics, Materials Innovation Factory, University of Liverpool, Liverpool, L7 3NY, UK; Centre of Excellence in Long acting Therapeutics (CELT), University of Liverpool, Liverpool, L69 3BX, UK

**Keywords:** COVID-19, SARS-CoV-2, Coronavirus, Pharmacokinetics, intracellular, active metabolite

## Abstract

**Background:** The role of favipiravir as a treatment for COVID-19 is unclear, with discrepant activity against SARS-CoV-2 *in vitro*, concerns about teratogenicity and pill burden, and an unknown optimal dose. In Vero-E6 cells, high concentrations are needed to inhibit SARS-CoV-2 replication. The purpose of this analysis was to use available data to simulate intracellular pharmacokinetics of favipiravir ribofuranosyl-5⍰-triphosphate (FAVI-RTP) to better understand the putative applicability as a COVID-19 intervention.

**Methods:** Previously published *in vitro* data for the intracellular production and elimination of FAVI- RTP in MDCK cells incubated with parent favipiravir was fitted with a mathematical model to describe the time course of intracellular FAVI-RTP concentrations as a function of incubation concentration of parent favipiravir. Parameter estimates from this model fitting were then combined with a previously published population PK model for the plasma exposure of parent favipiravir in Chinese patients with severe influenza (the modelled free plasma concentration of favipiravir substituting for *in vitro* incubation concentration) to predict the human intracellular FAVI-RTP pharmacokinetics.

**Results:** *In vitro* FAVI-RTP data was adequately described as a function of *in vitro* incubation media concentrations of parent favipiravir with an empirical model, noting that the model simplifies and consolidates various processes and is used under various assumptions and within certain limits. Parameter estimates from the fittings to *in vitro* data predict a flatter dynamic range of peak to trough for intracellular FAVI-RTP when driven by a predicted free plasma concentration profile.

**Conclusion:** This modelling approach has several important limitations that are discussed in the main text of the manuscript. However, the simulations indicate that despite rapid clearance of the parent drug from plasma, sufficient intracellular FAVI-RTP may be maintained across the dosing interval because of its long intracellular half-life. Population average intracellular FAVI-RTP concentrations are estimated to maintain the Km for the SARS-CoV-2 polymerase for 3 days following 800 mg BID dosing and 9 days following 1200 mg BID dosing after a 1600 mg BID loading dose on day 1. Further evaluation of favipiravir as part of antiviral combinations for SARS-CoV-2 is warranted.

## Introduction

The urgent global public health threat posed by COVID-19 has led the global scientific community to rigorously explore opportunities for repurposing existing medicines based upon either demonstration of auspicious antiviral activity against SARS-CoV-2, or a plausible mechanistic basis for anti-inflammatory / immunomodulatory activity. If sufficiently potent antiviral agents can be identified, there is potentially significant opportunity for deployment either as prophylaxis or in early infection to prevent development of severe disease. The role of antivirals in later stages of COVID-19 is less clear but cannot be rigorously assessed until sufficiently potent antiviral drug combinations become available. Several groups have highlighted the importance of considering the fundamental principles of clinical pharmacology when selecting candidates for investigation as antiviral agents [1-3], including a nuanced understanding of the principles of plasma protein binding [4]. For most successful antiviral drugs developed to date, a key principle is that plasma drug concentrations be maintained above *in vitro*-defined target concentrations (EC_90_ or protein binding adjusted EC_90_ where available) for the duration of the dosing interval. However, where the drug target resides intracellularly, and generation of an intracellular active metabolite is a prerequisite to unmask the pharmacophore, a more thorough understanding of the intracellular pharmacokinetics is required to rationalise doses required to maintain antiviral activity. For example, nucleoside-based drugs or prodrugs require intracellular phosphorylation by a cascade of host proteins, to generate tri- phosphorylated metabolites that exert the activity on the viral polymerase [5, 6]. Indeed, antiviral nucleoside analogues for HIV (e.g. tenofovir alafenamide), HCV (sofosbuvir) and several of the repurposing opportunities for SARS-CoV-2 (e.g. remdesivir) have active triphosphate metabolites with intracellular half-lives much longer than the parent drug in plasma [7-9]. Therefore, for many nucleosides being explored for SARS-CoV-2 (including favipiravir, molnupiravir), the activity may be maintained across the dosing interval despite plasma concentrations of the parent dropping below the EC_90_ at trough concentration (C_trough_).

Favipiravir is approved for influenza in Japan but not elsewhere and has been intensively studied as a potential antiviral intervention for several other RNA viruses; most recently SARS-CoV-2. For several reasons, considerable uncertainty exists about the suitability of favipiravir as a COVID-19 intervention. Concerns about teratogenicity and high pill burden may limit widespread uptake of the drug during early infection, particularly in the absence of concomitant contraceptive use in women of child-bearing age [10]. Furthermore, *in vitro* studies of favipiravir in Vero-E6 cell infected with SARS-CoV-2 have yielded inconsistent findings[11-14], and low potency (EC_90_ = 159 μM; 24.9 μg/mL) has been described in those studies that have shown activity [14]. Favipiravir plasma concentrations also appear to diminish, the longer that patients receive the medicine [10], and studies in severe COVID-19 disease have shown that plasma exposures are almost entirely abolished [15]. Despite the uncertainty and potential limitations, favipiravir has been demonstrated to exert antiviral activity against SARS-CoV-2 in the Syrian Golden Hamster model [16], and despite extremely high doses (1000 mg/kg intraperitoneally), C_trough_ values in this model were similar to those achieved in human studies of influenza (4.4 μg/ml in hamster versus 3.8 μg/ml day 10 trough in human; [16, 17]). Cell- free models have also demonstrated the ability of the intracellular ribofuranosyl-51-triphosphate metabolite (FAVI-RTP) to directly inhibit the SARS-CoV-2 polymerase [18].

The purpose of this work was to model from published plasma pharmacokinetic profiles, the likely concentrations of the intracellular active moiety (FAVI-RTP) and evaluate whether putative target concentrations necessary to inhibit SARS-CoV-2 can be pharmacologically attained in humans.

## Methods

### Prior In-Vitro Data

Data for the intracellular formation and catabolism of intracellular favipiravir-RTP (FAVI-RTP) in MDCK cells following incubation with parent favipiravir were digitised from previously published work [19]. Briefly, the *in vitro* experiments carried out by Smee et al. involved incubation of confluent layers of MDCK cells in T-25 flasks with media containing favipiravir at 32, 100, 320 or 1000 μM for varying durations. Smee et al. included 10% foetal bovine serum in media and plasma protein binding of favipiravir is relatively low in humans (54%; [10]). Therefore, given that the protein concentration in the media was low in these incubations, it was assumed that the nominal *in vitro* media concentrations of favipiravir for the incubations equated to the free drug concentrations.

At given timepoints, medium was removed, and cells were washed, lysed and FAVI-RTP was quantified in the lysate using HPLC with UV detection. For catabolism/elimination experiments MDCK cells were incubated with favipiravir containing media at the specified concentrations for 24h to allow production and accumulation of FAVI-RTP, before the media was removed and replaced with favipiravir-free media. Incubation then continued for a series of timepoints at which media was removed, and cell lysate was assayed for FAVI-RTP. Smee et al. present their FAVI-RTP quantification in normalised units of pmol/10^6^ cells according to the cell counts of the incubations, which was taken to represent the normalised intracellular *amount* of FAVI-RTP. In this work a value provided by Bitterman et al. [20] for the volume of an MDCK cell as 2.08 pL was then used to convert the FAVI- RTP quantification of Smee et al. to units of pmol/(10^6^ pL) = pmol/μL = μM.

### Modelling of In-Vitro Data

The data for intracellular production and elimination of FAVI-RTP were fitted with a mathematical model in the R programming environment (v 4.0.3) [21] to describe and parameterise the observed data as a function of the incubation media concentrations and time. This fitting made use of the Pracma library [22] and lsqnonlin function for nonlinear regression.

Data for intracellular production (in presence of favipiravir containing media) and elimination of FAVI-RTP elimination (on removal of favipiravir containing media after a 24h incubation and replacement with blank media) were combined into single time courses for each medium concentration, to then be described with the following ordinary differential (rate) equation mathematical model:

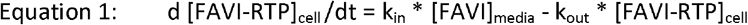

Where [] denotes concentration. The initial condition for [FAVI-RTP]_cell_ is set to zero at time zero and [FAVI] _media_ switched to equal zero at the 24h timepoint to end the production rate of FAVI-RTP and allow only the elimination rate to describe observed, declining concentrations from that time forward. The parameter k_in_ has units of time^-1^ and simultaneously describes net influx/diffusion of parent favipiravir from media into the cell and its (net) subsequent conversion to FAVI-RTP. The use of k_in_ therefore simplifies a more detailed description of favipiravir net influx into cells and conversion into FAVI-RTP into an empirical 1^st^ order process dependent on media concentration of favipiravir, to enable use of the information in the available data, where parent favipiravir itself is not quantified.

Smee et al. do not explicitly quote the volume of media used in their incubations, but given the T-25 flasks used, a media volume of 5-10 ml could be expected. Across the lowest to highest favipiravir media concentrations in the data (32 and 1000 μM), this therefore translates to a range of 0.16 to 10 μmol favipiravir present in the incubations at time = 0. The maximum amount of intracellular FAVI- RTP produced after 24h in the 1000 μM incubation was 332 pmol / 10^6^ cells ; with incubations declared to contain approximately 7 x 10^6^ cells on average, this gives a maximum total amount of 2,324 pmol = 2.3 nmol FAVI-RTP converted from favipiravir in 1:1 stoichiometry, which is << 1 % conversion of the total amount of favipiravir available in the 1000 μM incubation at the start (with similar calculations demonstrable at the other media concentrations). Therefore the [FAVI] _media_ term in Equation 1 can be considered approximately constant for a given time course dataset of incubations at a specified favipiravir concentration. In turn, this renders Equation 1 equivalent to a zero order constant input model with first order elimination and with the zero order input being switched off at 24h when media containing favipiravir was replaced with blank media.

### In-vivo Intracellular Simulations

The model and parameter estimates from the fitting to *in vitro* intracellular data, describing intracellular FAVI-RTP concentrations as a function of the media incubation concentrations were then taken forward and combined with a population PK model for plasma exposure for favipiravir described by Wang et al. in a Chinese population receiving the drug for influenza [17], substituting the media incubation concentration driving the intracellular FAVI-RTP production rate with the free plasma concentration predicted by the population PK model. This provided a prospective simulation of *in vivo* intracellular concentrations of FAVI-RTP (assuming cells of similar disposition to MDCK cells) as a function of *in vivo* plasma exposure.

The population PK model of Wang et. al is a 1-compartment PK disposition model with 1^st^ order absorption. Therefore, the equations for the model for *in vivo* intracellular simulations were as follows:

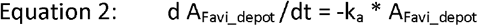

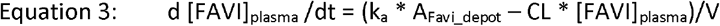

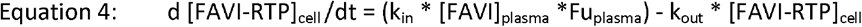

Where [] denotes concentration, CL and V are apparent values CL/F and V/F, and k_in_ and k_out_ used values derived from the *in vitro* model fitting. The Wang et al. model also incorporated a time dependent effect on CL, representative of favipiravir autoinduction of its own elimination, where:

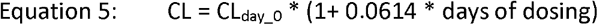

The model assumes a minimal proportion of the total mass balance of favipiravir transfers in from the plasma before conversion into the intracellular FAVI-RTP (similar to how the *in vitro* model assumes a constant [FAVI] _media_) and was therefore similar in some respects to a PKPD effect compartment model.

PK parameter population interindividual variabilities estimated by Wang et al. were used to simulate a population of 1000 sets of PK parameters and their resultant predicted [FAVI]_plasma_ and [FAVI- RTP]_cell_ profiles, from which 90% prediction interval profile envelopes were calculated. No population distribution of body weight was incorporated into this simulation which is equivalent therefore to assuming each simulated subject had a body weight of 70kg according to the Wang et al. population PK model. No interindividual variability in the k_in_ or k_out_ parameters was available or assumed. Parameter values used, and their inter-individual variabilities, quoted from the population PK model of Wang et al. are provided in Table 2, with Fu_plasma_ set at 0.46 [10].

**Table 2.** Summary of favipiravir POP-PK model parameter estimates used for simulation, from Wang et. al.

| Parameter | Est. | Inter-Individual Variance<br>($\omega^2$, log parameters) |
| --- | --- | --- |
| F<br>(apparent, fractional) | 1<br>(fixed) | 0.0921 |
| $k_a$<br>($h^{-1}$) | 1.5 | 1.05 |
| CL/F<br>(L/h) | 2.96 | 0.274 |
| V/F<br>(L) | 37.1 | 0.128 |
| Time dep. Eff. On CL/F<br>(% per day) | 6.14 | - |
| Interindividual Covariance<br>for $k_a$ and V/F<br>($\omega^2_{k_a} \sim \omega^2_{V/F}$) | - | 0.23 |

## Results

### Modelling of In-Vitro Data

Data from the observed [FAVI-RTP]_cell_ from Smee et al. at the four media concentrations investigated, overlaid with the model fittings are given in Figure 1, with parameter estimates and associated % relative standard errors in Table 1. The model was deemed to provide a satisfactory description of the observed data with acceptable precision of parameter estimates. However, it should be noted that k_in_ were not observed to be constant across the media incubation concentrations. A plot of k_in_ vs. [FAVI] _media_ (the latter on a log-scale) is provided in Figure 2, indicating that some form of saturable relationship is most likely present, which may require an E_max_ type model to be accurately described over the full range of media concentrations. However, with data available at only four media concentrations there was insufficient information to adequately fit such a saturable model. A log-linear model has therefore been fitted instead to the log-linear portion of the k_in_ vs. [FAVI] _media_ curve (the [FAVI] _media_ range from 32 to 320 μM, which also encompasses a typical range of *in vivo* plasma concentrations under standard human dosing regimens) and is overlaid in Figure 2, where:

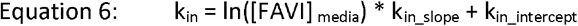

**Table 1.** Parameter estimates from *in vitro* PK model fittings

| [FAVI]<sub>media</sub> |  | k<sub>in</sub><br>(h<sup>-1</sup>) | k<sub>out</sub><br>(h<sup>-1</sup>) |
| --- | --- | --- | --- |
| 1000 µM | Est. | 0.020 | 0.126 |
|  | %RSE | 13.9 | 11.4 |
| 320 µM | Est. | 0.021 | 0.089 |
|  | %RSE | 14.7 | 15.6 |
| 100 µM | Est. | 0.033 | 0.112 |
|  | %RSE | 12.0 | 10.7 |
| 32 µM | Est. | 0.048 | 0.105 |
|  | %RSE | 15.3 | 14.3 |

**Figure 1.**
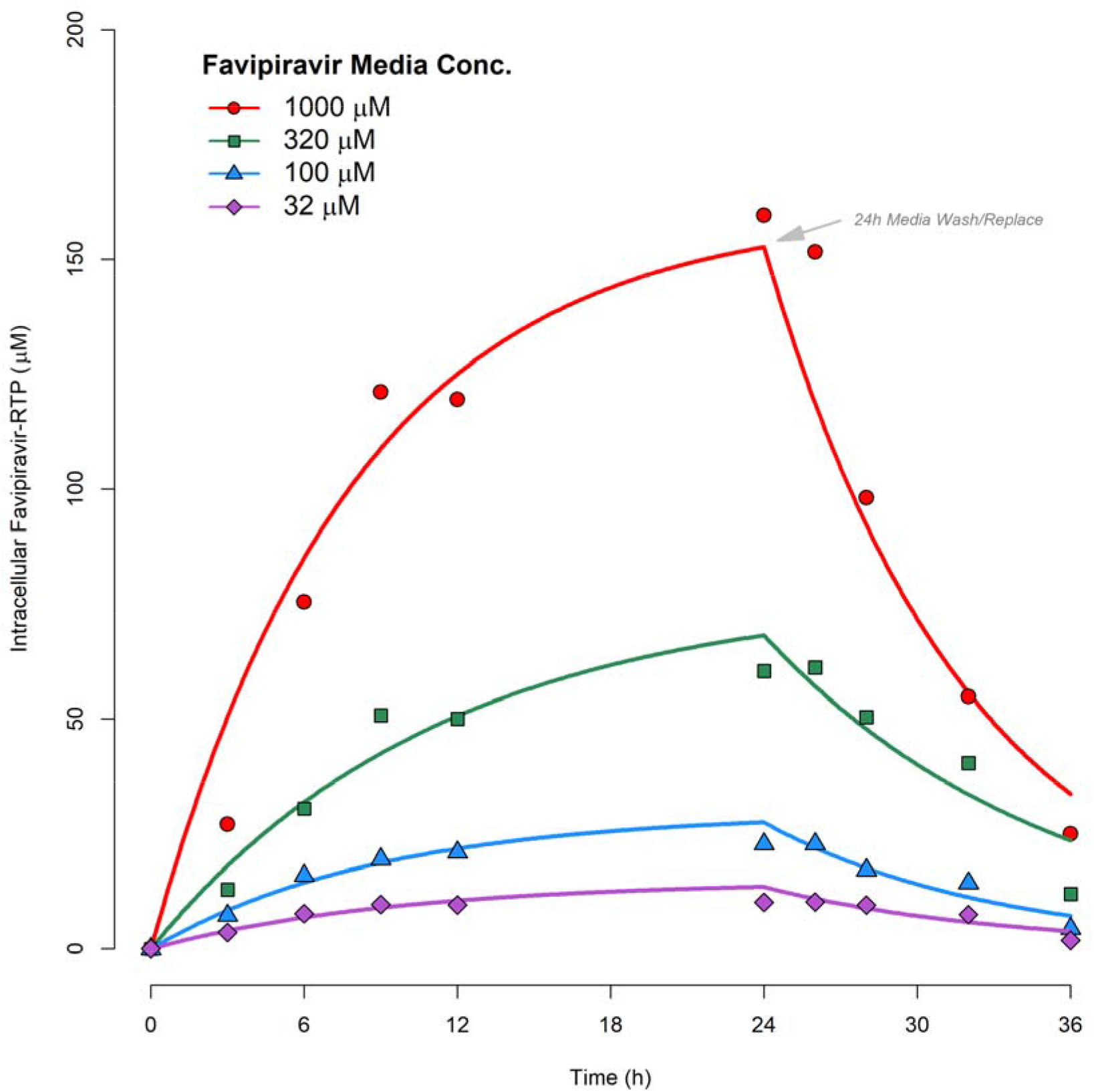
PK model fittings to time courses of [FAVI-RTP]_intracellular_ generated by Smee et al. in MDCK monolayers

**Figure 2.**
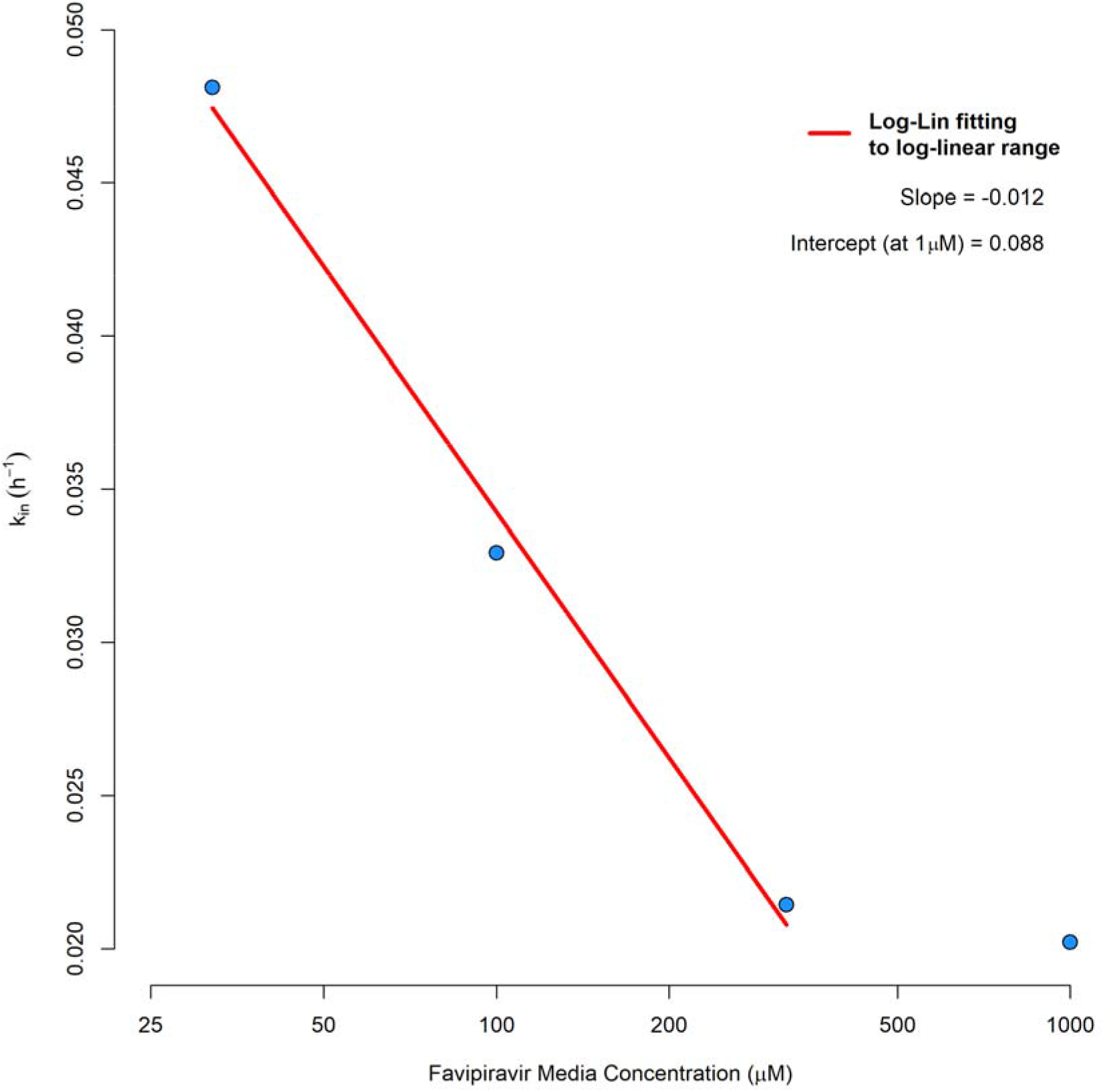
Log-linear fitting to k_in_ as a function media favipiravir concentration

The values of k_in_slope_ and k_in_intercept_ were then taken forward to the *in vivo* simulations instead of a mean value of k_in_, or the value from one [FAVI] _media_ fitting alone, and were used via Equation 6 during simulations to calculate the value of k_in_ required for any free plasma concentration at any given time as an input parameter value for Equation 4. k_out_ estimates from fittings were consistent at the 4 *in vitro* [FAVI] media concentrations. Therefore, the mean of the 4 estimates (0.108 h^-1^) was taken as the input value for simulations using Equation 4.

### In-vivo Intracellular Simulations

Simulations of predicted *in vivo* plasma and intracellular exposures for favipiravir and FAVI-RTP are shown in Figure 3A for a dosing regimen of 1600 mg BID loading dose for day 1, followed by 800 mg BID maintenance dosing for 9 further days. Simulations of predicted *in vivo* plasma and intracellular exposures for favipiravir and FAVI-RTP are shown in Figure 3B for a dosing regimen of 1600 mg BID loading dose for day 1, followed by 1200 mg BID maintenance dosing for 9 further days. In both cases, reference to the Km (Michaelis–Menten constant) for FAVI-RTP against the SARS-CoV-2 RNA-dependent RNA polymerase (RdRp) enzyme [18] is overlaid. The plasma target exposure based on *in vitro* EC_90_ for favipiravir of 159 μM against SARS-CoV-2 [14] is also shown but should be interpreted with caution owing to the lack of clarity in *in vitro* free drug concentrations and whether human plasma binding is high or low affinity.

**Figure 3.**
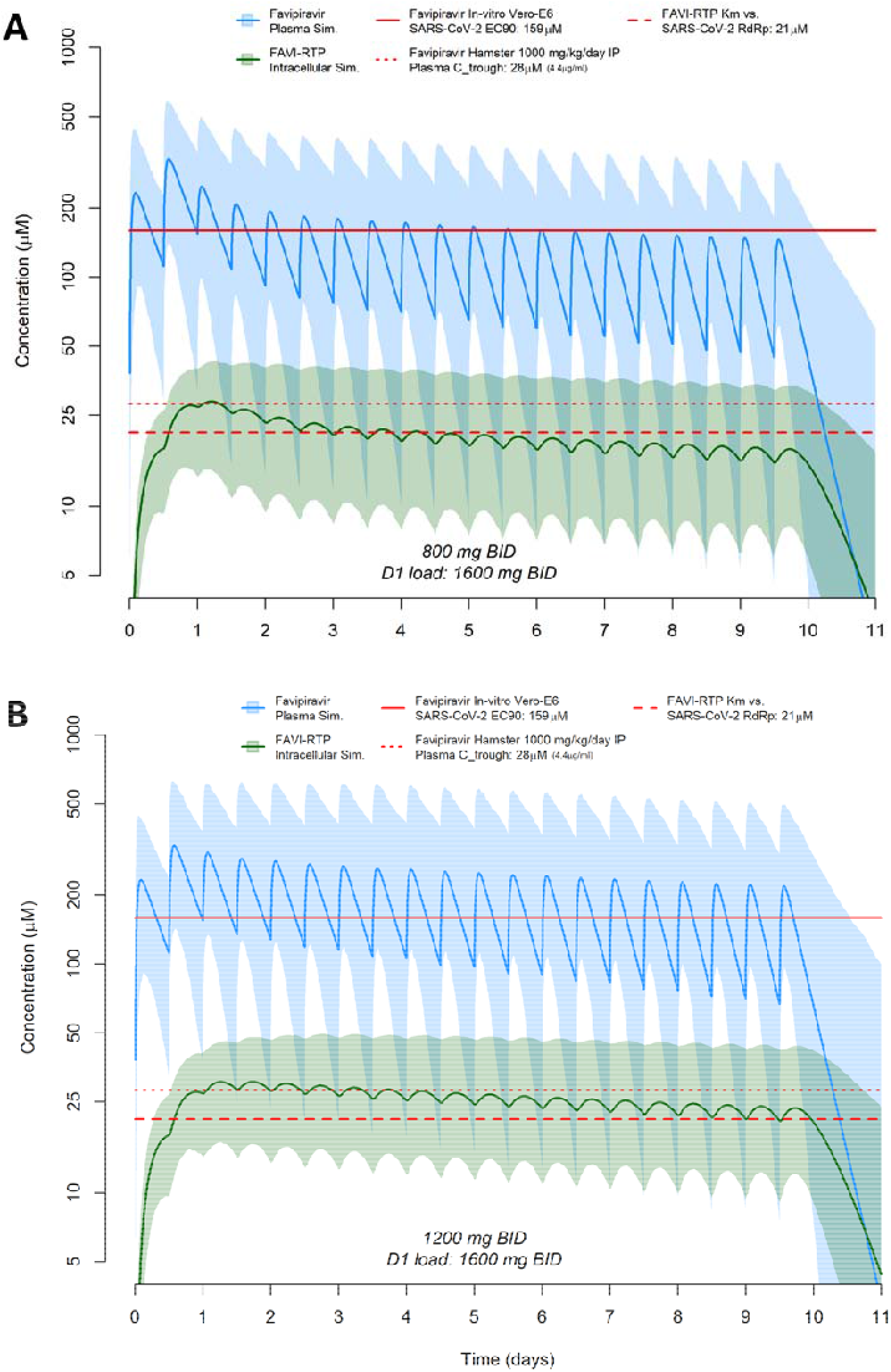
Favipiravir Plasma and Intracellular concentration predictions based on the Population-PK model of Wang et al. combined with *in-vitro* intracellular PK modelling, for a dosing regimen of 1600 mg BID loading dose for day 1, followed by 800 mg BID maintenance dosing for 9 further days (A), or 1600 mg BID loading dose for day 1, followed by 1200 mg BID maintenance dosing for 9 further days (B). Dashed red line represents the previously published Km for inhibition of SARS-CoV-2 polymerase by FAVI-RTP [18], dotted red line represents C_trough_ plasma concentrations of favipiravir following 1000 mg/kg/day dose in hamster [16] and solid red line represents the *in vitro* EC_90_ of favipiravir against SARS-CoV-2 in Vero-E6 cells [14].

## Discussion

While studies in the Syrian Golden Hamster have demonstrated effectiveness of favipiravir against SARS-CoV-2, *in vitro* activity data generated in the Vero-E6 cell model have questioned the utility of the molecule when the derived target concentrations are compared to the pharmacokinetics after administration to humans. Ultimately, robustly designed and executed clinical trials will be required to determine the utility of favipiravir for SARS-CoV-2 but understanding the mechanisms which underpin the clinical pharmacology is important to understand the plausibility for evaluation, which should underpin selection of candidates for clinical evaluation. Furthermore, an *a priori* understanding of likelihood of success for future candidates can only evolve from a thorough understanding of the PKPD rules of engagement for SARS-CoV-2, which currently do not exist. Antiviral drugs have only thus far been unequivocally successful for other viruses when given in combination, and the requirement of combinations to improve potency and/or stem emergence of resistance requires careful consideration so as not to obviate the lessons that should be learned from other pathogens. Notwithstanding, favipiravir has been evaluated at several different doses and schedules in numerous clinical trials globally, with mixed outcomes [23-25]. As of 29^th^ December 2020, a total of 44 trials were listed on clinicaltrials.gov aiming to evaluate favipiravir, predominantly as a monotherapy (with some exceptions) and in various use cases.

The current analysis aimed to apply a PK modelling approach to better understand the potential efficacy of favipiravir for SARS-CoV-2 at doses readily achievable in humans. The simulations synthesised available data for intracellular kinetics of FAVI-RTP in MDCK cells, plasma pharmacokinetics in a Chinese patient population, *in vitro*-derived antiviral activity data (EC_90_), and cell-free inhibition data for FAVI-RTP against the SARS-CoV-2 RNA polymerase. Importantly, this modelling approach indicates that despite rapid clearance of the parent drug from plasma, the peak to trough variability in intracellular FAVI-RTP is such that activity may be maintained across the dosing interval because of the long intracellular half-life. The simulations indicate that the population average intracellular FAVI-RTP concentrations will maintain its Km for the SARS-CoV-2 polymerase for 3 days following 800 mg BID dosing and 9 days following 1200 mg BID dosing after a 1600 mg BID loading dose on day 1. Importantly, the flatter intracellular pharmacokinetic profile of the phosphorylated form of favipiravir is in keeping with observations for other antiviral nucleoside/nucleotide analogues such as tenofovir-diphosphate [26, 27], which underpins the efficacy of these drugs for other viruses.

The current approach has several important limitations that should be recognised. Favipiravir pharmacokinetic exposures have been demonstrated to be lower in American and African patients than in Chinese patients [28], and so the simulations may not be widely applicable across different ethnicities. The modelling applied a direct *in vitro* to *in vivo* extrapolation of k_in_ and this should be considered as a major assumption as it directly presumes the *in vitro* Favi_media_ concentration is representative of free plasma concentration as derived from the PK model and that the umbrella k_in_ parameter, which consolidates various underlying uptake and conversion processes, is directly translatable. Importantly, the presented intracellular predictions are specific to data generated on intracellular kinetics in MDCK cells. Therefore, accuracy of the intracellular FAVI-RTP concentrations will be dependent upon the similarity of relevant human *in vivo* cells in terms of the *in vitro* uptake/elimination as well as the rate and extent of metabolic activation of favipiravir to its triphosphorylated active form. Furthermore, there are no data with which to model the inter-patient variability in the intracellular uptake or conversion to FAVI-RTP and so intracellular concentration variability shown in Figure 3 is only derived from intracellular variability in plasma exposure. Finally, the intracellular prediction is driven by the estimated free plasma concentration, whereas *in vivo* it is possible local tissue free drug concentrations at the target organ for which there are no available data may be higher or lower than in plasma.

Despite these limitations, additional confidence in the predictions come from two important sources. Firstly, following the first day of dosing, there is generally good agreement between the point at which the plasma favipiravir concentrations intersect the *in vitro* derived EC_90_ and the corresponding intracellular FAVI-RTP value being close to its Km value derived separately in a cell-free system with the SARS-CoV-2 polymerase (Figure 3). It should be noted that no data are available with which to derive a protein-adjusted EC_90_ value for favipiravir. Secondly, the clear activity of favipiravir in the Syrian Golden Hamster model when C_trough_ values are similar to those in humans gives additional confidence in the predictions [16].

In summary, these simulations indicate that favipiravir maintenance doses between 800 mg and 1200 mg BID may be sufficient to provide therapeutic concentrations of the intracellular FAVI-RTP metabolite across the dosing interval. Further evaluation of favipiravir as an antiviral for SARS-CoV-2 appears to be warranted and will provide additional clarity on its putative utility. However, the recent emergence of variants of the virus requires careful consideration of the drug resistance threat posed by using repurposed agents as monotherapies, particularly when they are likely not to be fully active in all patients. The polymerase, along with the protease and spike are likely to be extremely important drug targets for new chemical entities and care should be taken not to compromise their utility before the first-generation specific SARS-CoV-2 antivirals emerge.

## Data Availability

Data are available upon request.

## Acknowledgements

The Authors wish to thank Professor Leon Aarons (University of Manchester) for helpful discussions during development of this work.

